# The spatial and temporal distribution of SARS-CoV-2 from the built environment of COVID-19 patient rooms: A multicentre prospective study

**DOI:** 10.1101/2022.11.23.22282241

**Authors:** Michael Fralick, Madison Burella, Aaron Hinz, Hebah S. Mejbel, David S. Guttman, Lydia Xing, Jason Moggridge, John Lapp, Alex Wong, Caroline Nott, Nicole Harris-Linton, Rees Kassen, Derek MacFadden

**Author notes:** Correspondence, 60 Murray St, M5T3L9, Toronto, ON. **Study concept and design:** All authors. **Acquisition of data:** All authors. **Analysis/interpretation of data**: All authors. **Drafting of the manuscript:** Fralick M, MacFadden D. **Critical revision of the manuscript:** All authors. **Statistical analysis:** All authors. **Obtained funding:** This study was funded by the Sinai Health Department of Medicine Research Fund.

## Abstract

**Background:** SARS-CoV-2 can be detected from the built environment (e.g., floors), but it is unknown how the viral burden changes over space and time surrounding an infected patient. Characterising these data can help advance our understanding and interpretation of surface swabs from the built environment.

**Methods:** We conducted a prospective study at two hospitals in Ontario, Canada between January 19, 2022 and February 11, 2022. We performed serial floor sampling for SARS-CoV-2 in rooms of patients newly hospitalized with COVID-19 in the past 48 hours. We sampled the floor twice daily until the occupant moved to another room, was discharged, or 96 hours had elapsed. Floor sampling locations included: 1m from the hospital bed, 2m from the hospital bed, and at the room’s threshold to the hallway (typically 3 - 5m from the hospital bed). The samples were analyzed for the presence of SARS-CoV-2 using qPCR. We calculated the sensitivity of detecting SARS-CoV-2 in a patient with COVID-19, and we evaluated how the percentage of positive swabs and the cycle threshold of the swabs changed over time. We also compared the cycle threshold between the two hospitals.

**Results:** Over the 6-week study period we collected 164 floor swabs from the rooms of 13 patients. The overall percentage of swabs positive for SARS-CoV-2 was 93% and the median cycle threshold (for positive swabs) was 33.7 (IQR: 30.9, 37.5). On day 0 of swabbing the percentage of swabs positive for SARS-CoV-2 was 81.1% and the median cycle threshold was 33.7 (IQR: 32.1, 38.3) compared to swabs performed on day 2 or later where the percentage of swabs positive for SARS-CoV-2 was 98.1% and the cycle threshold was 33.4 (IQR: 30.7, 35.7). We found that viral detection did not change with increasing time (since the first sample collection) over the sampling period, OR 1.65 per day (95% CI 0.68, 4.02; p = 0.27). Similarly, viral detection did not change with increasing distance from the patient’s bed (1m, 2m, or 3m), OR 0.85 per metre (95% CI 0.38, 1.88; p = 0.69). The cycle threshold was lower (e.g. more virus) in The Ottawa Hospital (median Cq 30.8) where the floors are cleaned once daily rather than the Toronto hospital (median Cq 37.3) where floors were cleaned twice daily.

**Conclusions:** We were able to detect SARS-CoV-2 on the floors of rooms of patients with COVID-19 and the viral burden did not vary over time or by distance from the bed. These results suggest floor swabbing for the detection of SARS-CoV-2 in a built environment such as a hospital room is both accurate and robust to variation in sampling location and duration of occupancy.

## INTRODUCTION

SARS-CoV-2 primarily spreads via aerosols and droplets, and the degree of aerosolization is related to multiple factors including ventilation.^1–4^ Within the built environment, the floor is the most common location within the built environment where the virus can be detected.^5–9^ Floors likely act as a “sink” and collect the droplets and aerosols produced from infected individuals when they eventually fall to the floor. Hinz et al. conducted one of the first studies to identify whether the SARS-CoV-2 virus can be detected from the built environment within a hospital. They conducted a multicentre prospective study at two hospitals in Ottawa, Ontario, Canada where high touch surfaces (e.g., computer keyboard, door handle, telephone receiver, and various equipment) and the floors were swabbed weekly for a total of ten weeks.

They were able to recover viral RNA from these surfaces on wards dedicated for patients with COVID-19, but not on wards where there were no patients with COVID-19. The floor was the most common surface where the virus was detected and this observation has been replicated in other studies.^6,8^ A limitation of the study by Hinz et al. was that they did not swab within patient rooms and instead only swabbed the hallways of wards and other common areas within the hospital.

Floor swabbing has proven to be an effective approach for detecting and monitoring SARS-CoV-2 in hospitals. One of the first studies swabbing inside the rooms of patients with COVID-19 was by Zhang et al. They collected over 2000 environmental swabs on inpatient wards including in common areas and in the rooms of patients with COVID-19. In the common areas the percentage of swabs positive for SARS COV-2 was 75% and slightly higher within the patient rooms (77%). Kim et al. conducted a study at four hospitals in Korea assessing both air and surface contamination as well as the impact of surface cleaning on the ability to detect SARS-CoV-2. They collected 330 swabs of which, 27% were positive for SARS-CoV-2. Following routine cleaning procedures, they were unable to detect the virus on the surfaces. Notably, all of the 52 air samples collected from patient rooms and anterooms in the four hospitals were negative for SARS-CoV-2.

An important question is how the viral burden changes over space and time surrounding a patient, and how different cleaning protocols affect the viral burden. One single centre study in the US swabbed within the rooms of patients with COVID-19 to determine how severity of illness and distance from the patient’s bed affected the recovery of SARS-CoV-2 from the built environment (e.g., floors and high touch surfaces).^10^ They included 111 unique patient-room pairs and conducted a median of 1.5 swabs per patient-room pair. The probability of detecting SARS-CoV-2 from the floor was approximately 80% and this did not vary over distance, but was higher for patients with more severe disease (e.g., requiring positive pressure ventilation).

Because they did not perform serial swabs each day within the patient’s room, it is unknown how the probability of detection would vary over time. Furthermore, because their study was single centred and observational they were unable to identify how different cleaning protocols affected their findings. Our objective was to conduct serial swabs at systematic distances and times to understand how the viral burden changes over space and time and how different cleaning protocols affect viral burden. In doing so we can validate the prior work and also expand the current state of knowledge.

## Methods

### Study Design

We conducted a multicentre prospective study at two tertiary care hospitals in Toronto and Ottawa, Ontario, Canada between January 19, 2022 and February 11, 2022. Both of the hospitals have a combination of single and multi-patient rooms, however we only recruited single rooms where patients were hospitalized for COVID-19 in the preceding 48 hours. We swabbed the floors twice daily (at 9:00 and 17:00), at three distances from the hospital bed: 1 metre (m), 2 m, and where the room connected with the hallway (typically 3 to 5 m from the bed of the patient). Patient rooms were fully cleaned and disinfected before and after each admission. At The Ottawa Hospital, the floors and bathrooms of patient rooms were cleaned once daily while occupied by a SARS-CoV-2-infected patient, while the floors and bathrooms at the Toronto hospital were cleaned twice daily. At The Ottawa Hospital, this study was conducted under an existing umbrella REB approval, and for Mount Sinai Hospital the research was deemed excluded from requiring institutional approval given that it does not involve human subjects work.

### Environmental Detection of SARS-CoV-2 by RT-qPCR

Trained research staff swabbed the floors, with each swab taking approximately 30 seconds of swabbing across a 2” × 2” area. Floors were sampled using the P-208 Environmental Surface Collection Prototype kit from DNA Genotek (provided in-kind). The kit includes a flocked swab and 2 mL of semi-lytic nucleic acid stabilization solution for post-collection swab immersion.

SARS-CoV-2 was detected by quantitative reverse-transcriptase polymerase chain reaction (RT-qPCR) of the viral N-gene from RNA extracted from the stabilization solution using the MagMAX Viral/Pathogen II (MVP II) Nucleic Acid Isolation Kit (Thermo Fisher Scientific, Waltham, MA). Our previous study provides in depth information regarding the validation of SARS-CoV-2 detection from built environment swabs.^5^ The qPCR results provided us with a quantification cycle (Cq) of detection for each positive swab; we estimated the number of viral copies present using the Cq values and a previously-determined standard curve.^5^ For our study, we considered a positive result to be a Cq less than 45, which is a common cut-off used for environmental surveillance of SARS-CoV-2.^11^

### Study Outcomes

We hypothesised that SARS-CoV-2 detection would decrease with increasing distance from the patient’s bed and decrease over time from admission. We quantified the percentage of floor swabs positive for SARS-CoV-2, as well as the number of viral copies recovered per positive swab, and how these changed over space (e.g., distance from the patient’s bed) and time in each room.

## Statistical Analysis

All statistical analyses were performed using the R language (v4.1.2)^12^ and all figures were created with the ‘ggplot2’ package (v3.3.6). We used descriptive statistics to compare swab results (e.g., positivity and number of viral copies) over space and time. We calculated the sensitivity of surface swabbing under the assumption that all swabs would detect SARS-CoV-2 in the area immediately surrounding the patient. Confidence intervals for sensitivity estimates were computed using the Agresti-Coull method for binomial proportions (using the ‘binom’ package v1.1). Regretfully, 17 samples were lost or spoiled after collection and could not be tested; these observations were treated as missing at random in our analyses.

To examine differences in SARS-CoV-2 detection between hospitals, we first computed the room-level means for the proportion of positive swabs. Similarly, we computed the room-level (geometric) means for the number of viral copies using the log_10_ transformed values to reduce positive skew. We performed two-tailed student’s t-tests to compare hospitals using the room-level means for each outcome to avoid pseudoreplication.

We examined the effects of time and distance on the detection of SARS-CoV-2 using hierarchical mixed-effects models. In each model, random intercepts were included to account for correlation in the data due to repeated observations within rooms and the clustering of rooms within hospitals. A mixed-effects logistic regression model was created with SARS-CoV-2 detection as a binomial outcome with logit link function, where model parameters were estimated by maximum-likelihood using the Laplace approximation and Nelder-Mead optimization (using ‘glmer’ from ‘lme4’ v1.1). For fixed effects, we estimated odds ratios and 95% confidence intervals (Wald score method). Model fit was assessed by examining the residuals, fitted values, and dispersion. We left the values for time or distance unstandardized, such that their effect sizes could be interpreted in terms of days or metres, respectively. We used the unstructured default variance-covariance matrix for lme4.

We used the number of viral copies as a numeric outcome for a linear mixed-effects model to examine the effects of time and distance on the quantity of SARS-CoV-2 recovered from positive surfaces. Random intercepts were included for rooms clustered within hospitals. This model was created using the ‘lmer’ function from the ‘lme4’, with restricted maximum likelihood estimation and an unstructured covariance matrix.

## RESULTS

### Overall Findings

Over the 6-week study period, we collected 164 floor swabs from the rooms of 13 patients. The overall percentage of swabs positive for SARS-CoV-2 was 93% and the median cycle threshold (for positive swabs) was 33.7 (IQR: 30.9, 37.5) (Table 1). Overall, the median patient-room observation period lasted 48 h. However, patients tended to drop out earlier at the Toronto hospital, with a median observation period of 32 h, compared to 56 h at The Ottawa Hospital.

**Table 1.** Environmental detection of SARS-CoV-2 RNA in patient rooms.

|  | Patient rooms (n) | Samples (n) | Positives (n) | Mean Sensitivity (95% CI) | Median (IQR) log10 Copies |
| --- | --- | --- | --- | --- | --- |
| **Overall** |  |  |  |  |  |
|  | 13 | 164 | 152 | 0.93 (0.88, 0.96) | 2.08 (0.9, 2.88) |
| **Day 0** |  |  |  |  |  |
| Overall | 13 | 66 | 58 | 0.88 (0.78, 0.94) | 2.02 (0.6, 2.58) |
| 1 m | 13 | 23 | 21 | 0.91 (0.72, 0.99) | 2 (0.76, 2.43) |
| 2 m | 13 | 24 | 21 | 0.88 (0.68, 0.96) | 2.05 (0.44, 2.68) |
| 3 m | 11 | 19 | 16 | 0.84 (0.62, 0.95) | 1.98 (0.73, 2.3) |
| **Day 1** |  |  |  |  |  |
| Overall | 10 | 46 | 43 | 0.93 (0.82, 0.98) | 2.32 (0.79, 2.97) |
| 1 m | 10 | 17 | 16 | 0.94 (0.71, 1) | 2.57 (0.83, 2.96) |
| 2 m | 8 | 14 | 13 | 0.93 (0.66, 1) | 1.21 (0.76, 3.11) |
| 3 m | 9 | 15 | 14 | 0.93 (0.68, 1) | 2.58 (0.82, 2.79) |
| **Day 2** |  |  |  |  |  |
| Overall | 7 | 35 | 34 | 0.97 (0.84, 1) | 2.27 (1.48, 3.09) |
| 1 m | 7 | 13 | 13 | 1 (0.73, 1) | 2.25 (1.53, 2.57) |
| 2 m | 7 | 10 | 9 | 0.9 (0.57, 1) | 2.22 (1.46, 3.1) |
| 3 m | 7 | 12 | 12 | 1 (0.72, 1) | 2.52 (1.52, 3.66) |
| **Day 3** |  |  |  |  |  |
| Overall | 3 | 17 | 17 | 1 (0.78, 1) | 1.53 (1.03, 2.37) |
| 1 m | 3 | 5 | 5 | 1 (0.51, 1) | 1.36 (1.03, 1.54) |
| 2 m | 3 | 5 | 5 | 1 (0.51, 1) | 2.21 (1.52, 2.42) |
| 3 m | 3 | 7 | 7 | 1 (0.6, 1) | 1.53 (0.78, 2.38) |
| Footnote: IQR, interquartile range. CI, confidence interval |  |  |  |  |  |

Rooms where patients stayed longer generally had slightly greater SARS-CoV-2 detection in terms of sensitivity (0.09 ± 0.03 per day, F = 2.7, p = 0.02), but the number of copies recovered did not change significantly with the duration of the patient’s stay (0.42 ± 0.29 per day, F = 1.5, p = 0.17).

### SARS-CoV-2 Viral Detection Over Time and Distance

We created a mixed-effects logistic regression model to evaluate the effects of time and distance on SARS-CoV-2 viral detection in patient rooms, with random intercepts specified for rooms clustered within hospitals. We found that viral detection did not change with increasing time (since the first sample collection) over the sampling period, OR 1.65 per day (0.68, 4.02; p = 0.27). Similarly, viral detection did not change with increasing distance from the patient’s bed (1m, 2m, or 3m), OR 0.85 per metre (0.38, 1.88; p = 0.69) (Figure 1). The variance of the fixed effects time and distance (0.27) was very small compared to the variance of the random effects associated with hospital and room (7.69); while the residual variance was 3.29. The variance of the random intercepts for rooms (0.73; 9.5% of random effects variance) was very small compared to the variance for hospitals (6.96; 90.5%). A linear mixed model with the number of viral copies recovered as a continuous outcome showed similarly null results for the effects of time (estimate 0.0572; 95% CI −0.045, 0.16) and distance (0.10; −0.01, 0.22), with random effects accounting for the majority of variance.

**Figure 1.**
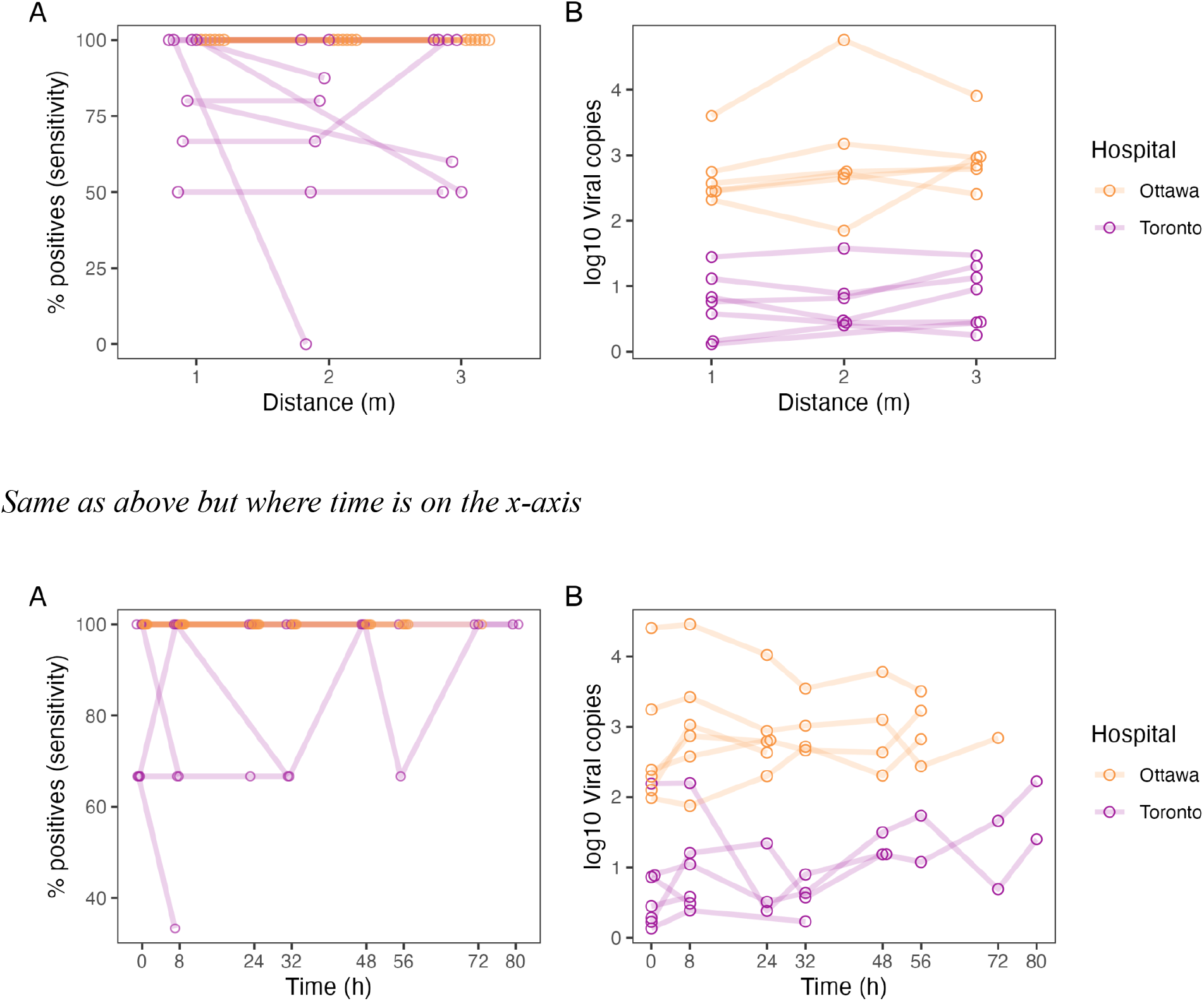
A comparison of % positivity [panel A] and estimated viral copies over space [panel B] Y-axis - % positive X-axis is distance from the hospital bed.

### Comparison Between Hospitals

We observed large differences in environmental SARS-CoV-2 detection in patient rooms at Ottawa and Toronto hospitals. At The Ottawa Hospital, 100% of samples were positive for SARS-CoV-2 for all rooms, whereas at the Toronto hospital the mean proportion of positives per room was only 78% (95% CI: 62-94%; p < 0.05). The number of viral copies quantified in positive samples was much greater for The Ottawa Hospital (794 copies; 95% CI: 171-3,017 copies) than the Toronto hospital (5.6 copies; 95% CI: 2.2 - 14 copies; p < 0.0001) (Figure 2).

**Figure 2.**
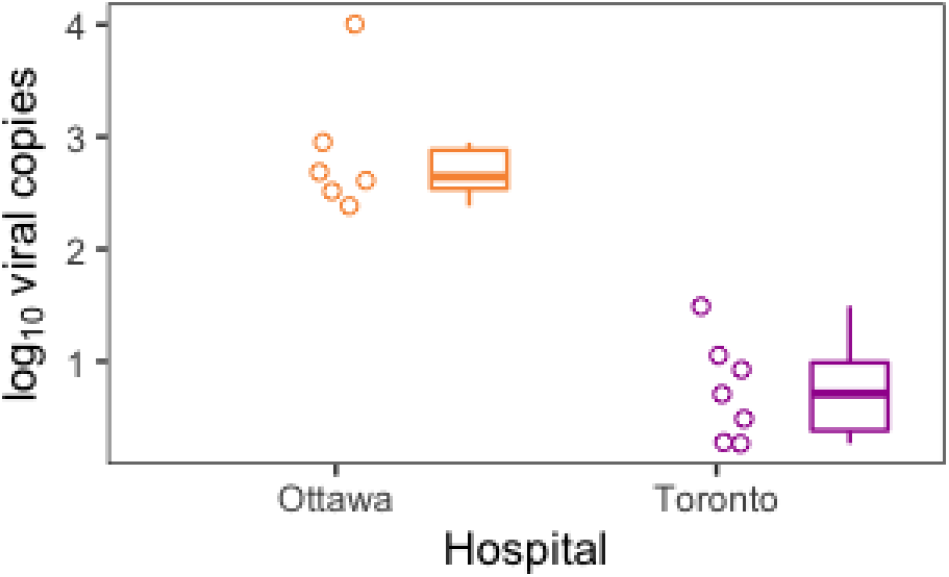
A comparison of detected SARS-CoV-2 quantities across two hospitals. Points show the patient means of log_10_ viral copies from all the positive swabs collected (negative results were excluded from the calculation of patient-level means). Box-plots summarise these values for each hospital.

## DISCUSSION

In this multicenter prospective study, we were able to identify and recover SARS-CoV-2 from the floors surrounding all of the included patients. The viral burden did not increase over time and the virus was consistently identified at 1 m and 2 m from the hospital bed, as well as at the entryway. The calculated sensitivity for swabbing the floor was 93%, indicating that floors serve as accurate indicators of the presence of patients with COVID-19.

Our findings align with the body of literature that SARS-CoV-2 can be detected from the floors of areas where there are patients with COVID-19. In our study, 100% of patients had at least one swab positive for SARS-CoV-2 on the first day of swabbing which demonstrates that the contamination of the floors occurs quickly. Patients were generally unmasked, so most expelled viral particles presumably end up on the floor. Our findings also suggest that frequent cleaning procedures may result in a lower burden of virus recovered, as SARS-CoV-2 RNA was detected more frequently at a hospital with once daily cleaning than at a hospital with twice daily cleaning (Figure 2). In our study we did not evaluate whether swabbing immediately after cleaning resulted in an inability to detect SARS-COV-2. However, prior studies have done so.^13–15^ For example, Ong et al. identified that following routine cleaning they were unable to detect SARS-CoV-2 using PCR compared to prior to cleaning the surface. In the study by Kim et al. RNA was not detected in a room routinely cleaned by disinfectant wipes demonstrating how cleaning removes SARS-CoV-2 from the surface. However, RNA was detected in a room sprayed with disinfectant, suggesting disinfectant sprays may not be effective in reducing exposure. These findings confirm that what is detected on the floor is not simply a reflection of prior patients in the room.

We also identified that the virus could be consistently identified at all distances from the patient’s bed where we swabbed (e.g., 1 m, 2 m, entryway) (Figure 3). It is important to note that the rooms we studied had patients who are typically confined to their beds because of their oxygen requirements from the severe fatigue and weakness of COVID-19. Thus, our ability to consistently detect the virus at increasing distances from the patient’s bed goes against the historically referenced “6 feet” rule.^16^ The six foot rule was recommended by the CDC and other agencies based on the assumption that COVID-19 is spread via large droplets that can only travel short distances and thus staying 6 feet apart could help prevent the spread of COVID-19. Our study adds to the available literature that the virus can rapidly reach distances beyond 6 feet.

**Figure 3.**
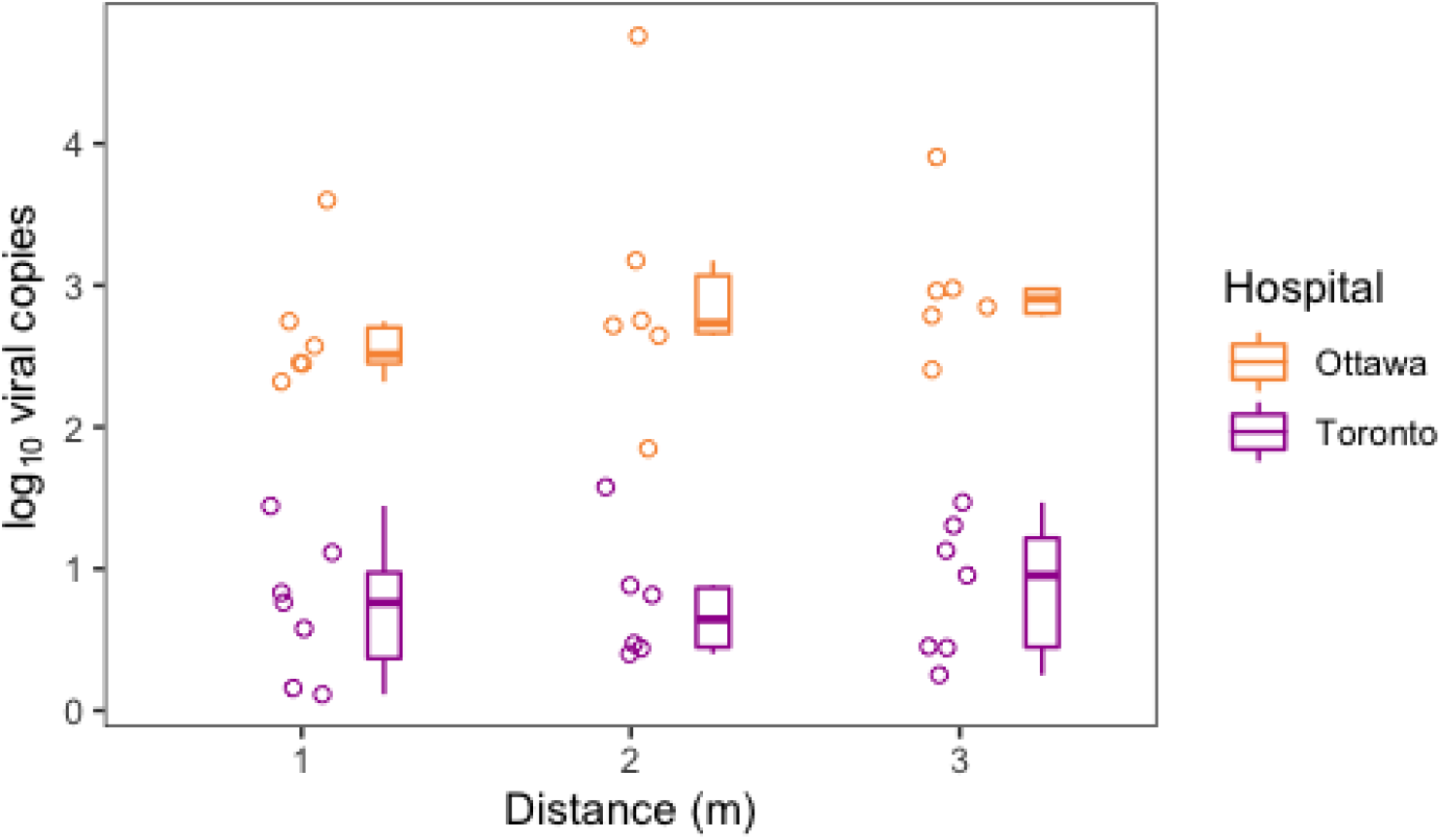
A comparison of detected SARS-COV-2 over distance across two hospitals. Points show the patient means of log_10_ viral copies from all the positive swabs collected (negative results were excluded from the calculation of patient-level means). Box-plots summarise these values for each hospital.

Our results highlight that swabbing floors for SARS-CoV-2 may be a practical tool for viral surveillance in settings where individual testing is not regularly performed.^5^ This environmental sampling technique may help identify locations of outbreaks, predict future outbreaks in advance of confirmed cases, and guide disinfection protocols in healthcare settings.^5^

There are other important limitations to our study. First, while we conducted 164 swabs, our study was relatively small as we only included 13 unique patients. However, our results align with prior studies and an added strength of our work is that we include two hospitals in different cities to improve generalizability. Second, we did not collect patient-level data to investigate how severity of illness or degree of immunocompromise may have influenced the degree of viral burden detected. However, a prior study observed higher rates of surface contamination with SARS-CoV-2 for patients who required high-flow oxygen or positive pressure ventilation compared to hospitalized patients with less severe illness (OR=1.6, 95% credible interval [CrI] 1.03-1.25).^10^ Third, by definition we only included rooms of patients with COVID-19 who were hospitalized on a medical ward and thus it is unknown whether our findings apply for patients with mild disease who did not require hospitalization. Finally, our study focused on SARS-CoV-2 and thus it is unknown how our results will apply to other respiratory pathogens such as influenza or respiratory syncytial virus. This will be an important area of future work and our study design can be easily adapted to study these, and other, pathogens.

## Data Availability

All data produced in the present study are available upon reasonable request to the authors

